# Rethinking intensity: Repetitive transcranial magnetic stimulation across ethnic populations

**DOI:** 10.1101/2025.02.22.25322728

**Authors:** Bella B B Zhang, Tim T Z Lin, Penny P Qin, Minxia Jin, Adam W L Xia, Rebecca L D Kan, Frank F Zhu, Georg S Kranz

## Abstract

Repetitive transcranial magnetic stimulation (rTMS) targeting the left dorsolateral prefrontal cortex is a widely used adjunctive therapy for various neurological and psychiatric disorders. Traditionally, the standard practice for dosing rTMS has involved adjusting the intensity to a defined percentage of the motor threshold. However, we perceive anatomical differences among ‘*White’*, ‘*Han Chinese*’ and ‘*Black or African American’* ethnic groups that result in unequal doses of stimulation at the left dorsolateral prefrontal cortex, a prime target for therapeutic rTMS. Therefore, we hypothesize that diverse ethnic populations experience different therapeutic effects with FDA-approved rTMS protocols that have been adopted globally. This also raises concerns about varying therapeutic effects and potential biases in existing research comparing efficacy across ethnicities. More importantly, our hypothesis emphasizes the need to tailor rTMS safety guideline for diverse ethnic populations. We call for well-powered clinical trials to systematically examine the effects of rTMS intensity in diverse ethnic groups.

## Introduction and hypothesis

Repetitive transcranial magnetic stimulation (rTMS) is a widely used adjunctive therapy for various neurological and psychiatric disorders including depression, fibromyalgia, Parkinson’s disease, addiction, and schizophrenia, among others.^1^ rTMS involves applying magnetic pulses to a specific brain region, with the left dorsolateral prefrontal cortex being a clinical prime target.^2,3^ For over three decades, the standard practice for dosing rTMS has involved adjusting the stimulation intensity at the target region to a defined percentage of the motor threshold, the minimum intensity needed to elicit a motor response at the motor cortex.^4,5^ Notably, US FDA-approved rTMS protocols generally use 120% of the motor threshold, which has been adopted globally.^6^ However, underlying this strategy is the assumption of a uniform relationship between the stimulation intensity and its therapeutic effects. That means we may have assumed that the ratio of scalp-to-cortex distance at the left dorsolateral prefrontal cortex to the motor cortex is identical across individuals. Considering anatomical differences such as head morphologies among diverse world populations,^7^ the authors see a need to re-examine this generic strategy for determining rTMS intensity, and hypothesize that the optimal intensity differs among ethnic groups.

## Contributing observation 1: ethnic differences in the scalp-to-cortex distance

We examined the scalp-to-cortex distances at the left dorsolateral prefrontal cortex and the left motor cortex among three ethnic groups (aged 22-35 years): 821 ‘*White*’, 165 ‘*Black or African American*’, and 159 ‘*Han Chinese*’ individuals. We obtained unprocessed T1-weighted and T2-weighted structural MRI images from of the Human Connectome Project (https://db.humanconnectome.org) and the Chinese Human Connectome Project (https://doi.org/10.11922/sciencedb.01374). We defined the locations of the left dorsolateral prefrontal cortex and the motor cortex as the 10-10 EEG positions F3 and C3, respectively.^8^ We used the SimNIBS package to segment the MRI images and created a head volume conductor model for each individual.^9^ The results were visually inspected for errors in tissue boundary establishment. We calculated the scalp-to-cortex distance as the Euclidean distance between the EEG position and the nearest site on the gray matter.

We found that the ‘*White’* individuals had a larger mean scalp-to-cortex distance at the left dorsolateral prefrontal cortex compared to the motor cortex (mean F3/C3 distance ratio=1.077). The relationship was reversed for the ‘*Han Chinese*’ individuals (mean F3/C3 distance ratio=0.939). The ‘*Black or African American*’ individuals had similar mean distances at the two cortices (mean F3/C3 distance ratio=0.992). An analysis of covariance, controlling for age and gender as covariates, revealed that the scalp-to-cortex distance ratio significantly differed among the three ethnic groups (F_2,1141_=75.580, p<0.001, ◻^2^=0.112). Post hoc pairwise comparisons showed that the ‘*White*’ individuals had a significantly larger ratio than both the ‘*Han Chinese*’ and the ‘*Black or African American*’ individuals (p’s < 0.001, Bonferroni corrected) (Figure. 1a).

**Figure 1.**
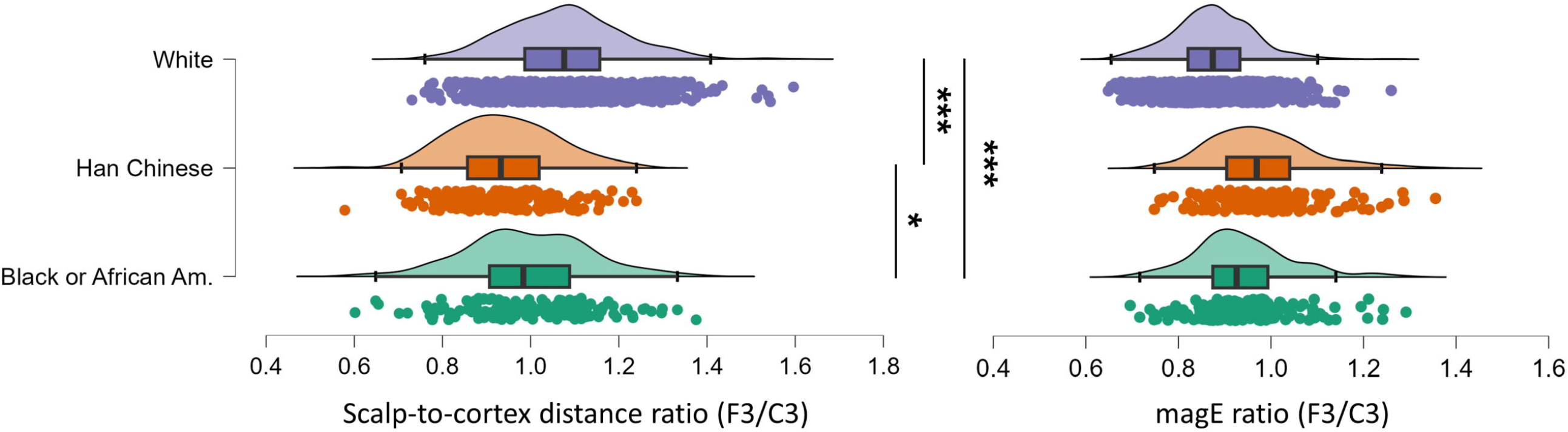
Scalp-to-cortex distance ratio and electric field strength ratio across ethnicities. Left: scalp-to-cortex distance ratio (F3/C3) for the left dorsolateral prefrontal cortex relative to the motor cortex in *‘White’, ‘Han Chinese’*, and *‘Black or African American’* individuals. A ratio greater than 1 indicates a larger scalp-to-cortex distance at the left dorsolateral prefrontal cortex compared to the motor cortex. Right: electric field strength ratio (F3/C3) for the left dorsolateral prefrontal cortex relative to the motor cortex in the three ethnic groups. A ratio below 1 indicates lower electric field strength in the left dorsolateral prefrontal cortex compared to the motor cortex. *: p<0.05, ***: p<0.001.

## Contributing observation 2: simulated optimal intensities for different ethnic groups

The marked differences in scalp-to-cortex distances led us to investigate the electric field strength induced by rTMS in the two cortices. We simulated the electric field strength for the three ethnic groups using SimNIBS 4.1.0.^10^ We assumed a standard rTMS treatment coil (MagVenture Cool-B65^11^) was positioned on the scalp at the EEG position F3 (left dorsolateral prefrontal cortex) and C3 (motor cortex) at a 45-degree angle to the midline, with the handle pointing posteriorly.^8^ We set the stimulation intensity to 1e6 A/s and calculated the induced electric field strength as the average value^12^ for every gray matter voxel within a 10-mm-radius sphere centered at the cortical projections from the EEG position F3 or C3.^13^ The F3 projection was at (x=-35.5, y=49.4, z=32.4 mm) and C3 at (x=-52.2, y=-16.4, z=57.8 mm) in Montreal Neurological Institute coordinates. An analysis of covariance, controlling for age and gender, indicated a significant effect of ethnicity on the electric field strength ratio of the left dorsolateral prefrontal cortex to the motor cortex (F_2,1141_=86.333, p<0.001, ◻^2^=0.128). While all groups exhibited an F3/C3 electric field strength ratio below 1, suggesting weaker stimulation received by the left dorsolateral prefrontal cortex compared to the motor cortex, the ratio differed significantly between the groups (p’s<0.001, Bonferroni corrected). On average, ‘*White*’ individuals received lower electric field strength in the left dorsolateral prefrontal cortex compared to the left motor cortex (F3/C3=0.875) when compared with ‘*Han Chinese*’ individuals (F3/C3=0.977) and ‘*Black or African American*’ individuals (F3/C3=0.940) (Figure. 1b). Specifically, if stimulation intensities at the left dorsolateral prefrontal cortex were set to the FDA-approved 120% of the motor threshold, then on average, ‘*Han Chinese*’ individuals would receive an equivalent intensity of 134% to stimulate ‘*White’* individuals, and ‘*Black or African American’* individuals would receive an equivalent intensity of 129% to stimulate ‘*White’* individuals.

## Implications of the hypothesis

Our analyses revealed significant differences in the scalp-to-cortex distances among the ‘*White’*, ‘*Han Chinese*’ and ‘*Black or African American’* ethnic groups. We further showed that the three ethnic groups would receive unequal doses of stimulation at the left dorsolateral prefrontal cortex, a prime target for therapeutic rTMS, even when the stimulation was applied at the same percentage of the motor threshold. Our findings align with previous empirical research and modeling studies. ^14,15^ Therefore, we hypothesize that the three ethnic groups experience varying clinical effects with standard FDA-approved rTMS protocols. Specifically, we presume that ‘*Han Chinese*’ and ‘*Black or African American*’ individuals have been suboptimally stimulated with standard rTMS stimulation protocols, considering that early rTMS clinical trials were predominantly conducted in North America and recruited disproportionately more ‘*White*’ individuals (thus, these protocols were presumably better optimized for the ‘*White*’ ethnic group).^16–19^ Our hypothesis further suggests that systematic biases may exist in previously published meta-analyses in terms of comparing therapeutic effects across ethnicities and summarizing effect size estimates. More importantly, our hypothesis highlights the need to tailor rTMS safety guidelines for different ethnic groups. Indeed, our simulation revealed that the FDA-approved stimulation intensity of 120% of the motor threshold, when applied to an average ‘*Han Chinese*’ individual, may generate an electric field strength equivalent to that produced by an intensity of 134% applied to an average ‘*White’* individual. Yet, rTMS at an intensity of 130% or above remains largely uncharted in research and is subject to a sharply reduced maximum safe train duration to prevent seizures.^20^

## Testing the hypothesis

We propose a series of rTMS clinical trials across the globe to test our hypothesis. We propose to start with trials on major depressive disorder, the disorder with the strongest evidence for the efficacy of rTMS on the dorsolateral prefrontal cortex. These trials will require coordinated planning and implementation across study sites to ensure a comparable treatment process. The first trial will focus on ‘*Han Chinese’* and ‘*White*’ patients, since this pair of ethnic groups demonstrated the largest difference in the scalp-to-cortex distance ratio. The standard, FDA-approved stimulation intensity of 120% of the motor threshold and the intensity of 108% adjusted for ‘*Han Chinese’* will be applied to patients from both ethnic groups. We expect that the more effective treatment intensity, in terms of response or remission rates, will vary between the two ethnic groups. We estimate that a few hundred patients per ethnic group will allow the detection of such clinically meaningful differences. If the results turn out to be positive, we may proceed with comparisons involving *‘Black or African American’* and ‘*White’* patients in a similar manner, though that would be a study on a much larger scale. Findings from this series of trials may give birth to a readily implementable strategy that benefits patients who cannot have magnetic resonance imaging scans for more precise structural measurements due to cost, availability, or safety considerations.

## Data Availability

All data produced in the present study are available upon reasonable request to the authors.

## Contributors

BBBZ: conceptualization, data curation, investigation, formal analysis, software, visualization, methodology, writing – original draft, writing – review & editing; TTZL: conceptualization, data curation, investigation, software, methodology, formal analysis, validation, writing – original draft, writing – review & editing. JMX: investigation, validation, writing – review & editing; AWLX: investigation, validation, writing – review & editing; PPQ: investigation, software, validation, writing – review & editing; RLDK: investigation, validation, writing – review & editing; FFZ: conceptualization, methodology, writing – original draft, writing – review & editing; GSK: conceptualization, writing – original draft, writing – review & editing, resources, supervision, project administration.

